# Relationships Among Knee Injuries, Q-angles, and Competition Levels in Female Ski Racers

**DOI:** 10.1101/2023.09.14.23295552

**Authors:** Danielle Taylor, Catherine Bevier

## Abstract

Prior research has focused solely on the prevalence of anterior cruciate ligament (ACL) injuries in male and female alpine ski racers and on the Q-angle, the angle created between the quadriceps muscles and the patella tendon, in ways unrelated to alpine ski racing. A survey was sent to female alpine ski racers who represent the academy U16 level through World Cup professionals that included questions regarding the athlete’s racing level, Q- angle, and past knee injuries. A total of 124 responses were collected. Just under half (45.16%) of the survey participants reported a history of at least one knee injury. A great majority, 83.33%, of the Professional athletes reported tearing their knee at least once, which is 3.04 and 1.90 times more than athletes of the Pre Professional and U16 levels respectively. Athletes with a Q-angle of 15° or 20° had torn 85.26% of the total ligaments with the ACL, MCL, and meniscus being the most frequently torn ligaments. From this self-reported data, it is clear that Q-angles and racing level both influence the frequency of ligament tears in female alpine ski racers. An athlete competing at a higher level of competitive ski racing is more likely to tear a knee ligament than those at lower levels, and larger Q- angles influence the risk of injury as well as injury to specific ligaments within the knee.

## Key Points

- Injury risk increase as ski racing level increases
- Injury risk increases with larger Q-angles

## Declarations

i. List of abbreviations-FIS; Fédération Internationale de Ski, NorAm; North American Cup, ACL; Anterior Cruciate Ligament, PCL; Posterior Cruciate ligament, MCL; Medial Collateral Ligament, LCL; Lateral Collateral Ligament, ANOVA; Analysis of variance, G’s; Gravitational forces.
ii. Ethics approval and consent to participate-Approved by the Internal Review Board at Colby College #2020-003. A virtual signature by legal guardians of all minors was obtained.
iii. Consent for publication-Not applicable.
iv. Availability of data and material-All data was submitted as supplementary material.
v. Competing interest – Not applicable.
vi. Funding - There was no funding received.
vii. Author contributions - DT is the sole author and contributor to this work. DT conducted all research activities including the experimental plane, design, data collection, analysis, and manuscript writing.
viii. Acknowledgements - I would like to thank Dr. Bevier for assisting with editing and Benefit Allocation Systems for donating the survey software. I would also like to acknowledge all athletes who participated in this study.
ix. Authors’ information-DT is a retired FIS (Fédération Internationale de Ski) athlete. She skied at the Pre Professional Level for 6 years and at the U16 Level for 2 years. Currently an Undergraduate student at Colby College pursuing Bachelor of Arts in Biological Science. DT is the soul author.

## 1. Introduction

### 1.1 Risks of alpine ski racing

Alpine ski racing is a high-risk sport with a high rate of knee injuries. The sport itself is unique because both genders compete and train under the same conditions. Females, however, are 2.3 times more likely to sustain a knee injury than their male counterparts [5]. The external risks that accompany alpine ski racing are virtually identical across the board for males and females. Some of these factors include types of slopes, conditions, amount of on-snow training, and dryland regimens. Conditions for athletes at the U16 academy level through World Cup level are also extremely similar, with conditions typically being slightly more dangerous at the World Cup level. The athletes within these three groups are some of the most elite female ski racers in the country in their respective levels. This ensures that similar training regimens have been in place for all of these athletes since their U14 years. This makes ski racing an optimal sport for assessing intrinsic and extrinsic factors that influence injuries.

### 1.2 Ski racing level

One factor that can influence knee injury is the level of racing competition. At the U16 level, where competitors are full-time student athletes (14-16 years old) the forces created, and the speeds reached on the course, are dramatically greater than those of lower levels of competition. These force and speed metrics tend to increase with each competition level. This makes crashes more frequent as the increased forces and speeds can be difficult to manage. For many FIS (Fédération Internationale de Ski) athletes, and specifically those competing in Entry Level FIS races, some competition and training venues are injected with water to save the integrity of the snow. But injecting the snow makes the slope extremely icy and hard-packed. Courses where Professional skiers compete, including venues for the North American Cup (NorAm), Europa Cup, and World Cup, are almost always injected. Injected snow is beneficial for performance and most athletes prefer this treatment. However, it creates more risks during crashes because athletes will continue sliding down the slick slope for a longer period of time and are able to reach higher speeds on this type of snow. This makes forces, such as gravitational forces, also greater. Since the forces created in the course are so large, weight training is required in all academy programs and above so that athletes can better manage the forces they will experience at higher levels of competition.

Ski racers at the U16 level as well as higher levels of competition all undergo daily weight training and other forms of conditioning to avoid injuries caused by weak muscles or ligaments. Even with daily training, young ski racers are still prone to knee injuries. For example, female athletes surveyed at age 19, who all started ski racing at age 9, on average experienced at least one tear of the anterior cruciate ligament (ACL) by age 17 [5]. Furthermore, once female athletes reach an age of about 16.7 years, the risk of injury is as high as it is at the Professional level [4]. This is the approximate age that females are fully mature in terms of growth, and therefore have larger Q-angles than adolescent girls or males. This is due to a girl’s hips widening during puberty which then leads to a larger Q-angle once they fully mature.

### 1.3 Q-angles

In general, females tend to have larger Q-angles because of their wider hips, so this leg posture may influence injury along with race course conditions. The Q-angle is defined as the angle between the Anterior Superior Iliac Spine and the central patella along an imaginary line drawn from the central patella to the tibial tubercle [7]. This distance tends to be greater in females than in males; the mean Q-angle for women was 15.8° in a supine position and 17.0° in a standing a position, which are both larger compared to the same metrics for men (12.7° and 13.6°, respectively) [8]. These differences make women more susceptible to knee ligaments injuries than their male counterparts. For women, a Q-angle greater than 20° is knock-kneed, a Q-angle less than 10° is bow-legged, and a Q-angle of about 15° is relatively straight [7]. These ligaments help support the articulation of the femur and tibia, which aid in forming the Q-angle. The knee support system is composed of many different ligaments, muscle, and cartilage that all play an important role in this articulation. The ligaments include the anterior cruciate ligament (ACL), posterior cruciate ligament (PCL), medial collateral ligament (MCL), lateral collateral ligament (LCL), patellar tendon, and meniscus.

### 1.4 Canting of ski boots

Alpine ski racers can compensate for the effect of their Q-angle on their skiing ability by having their boots canted. Canting is the process of manipulating the shell of the ski boot to aid in correcting the stance of a person with a wider or smaller Q-angle enabling them to stand flat on their skis. There are a few main ways to cant a boot, the most common way being to shave down one side of the shell of the boot to a specific degree. Canting strips can be used the same way as shaving a boot down, but they are usually not a permeate solution. Another more permeate solution that can be used in conjunction with the shaving of a boot is cuff rotation. This is when the cuff of the boot is tilted out or in to help position the tibia perpendicular to the ground. There are other forms of canting but these are the most common forms that boot fitters and coaches generally use. Canting takes into account that individual skiers have different builds and gives everyone equal ability to put their skis on edge despite anatomical differences. Canting adds or subtracts angle degrees on one side of the foot to compensate for the uneven stance created by the Q-angle, but it does not help to prevent knee injuries, because it does not change the Q-angle, it only changes the athlete’s stance on their skis.

### 1.5 Hypothesis

Extensive research is available that compares male and female knee injuries, and some studies focus on ACL-specific injuries in females. However, there are no published studies that focus on injury to multiple ligaments of the knee in female athletes (particularly alpine ski racers) and how anatomy can play a role in predicting injury probability. The goal of this study is to determine if different Q-angles in female alpine ski racers differentially affect which ligaments in the knee are more likely to be torn, as well as the frequency of ligament tears occurring at different levels of competition.

I propose two hypotheses to address this issue. First, the competition level of ski racing influences the likelihood of an athlete to tear a knee ligament. I expect that U16 ski racers exhibit lower frequencies of knee ligament injury than Professional ski racers. Second, different Q-angles influence the risk of injury to specific ligaments within the knee. I predict that a Q-angle of 20° leads an athlete to be more prone to injury of the MCL, ACL, and patellar tendon. An athlete with a Q-angle of 15° would be more prone to injury of the ACL and meniscus. Finally, an athlete with a Q-angle of 10° would be more prone to injury of the LCL. Comparing the Q-angle of the athletes with the ligaments they have torn, and their ski racing level will yield a comprehensive view of how different Q-angles and competition levels can influence injuries.

## 2. Methods

### 2.1 Survey

From January to September 2020, I sent out surveys to female U16s, Entry Level FIS, NorAm, Europa Cup, and World Cup alpine ski racers. I designed the survey using Alchemer. This survey received approval (#2020- 003) from the Internal Review Board at Colby College. The survey was anonymous and included questions regarding Q-angles, ski racing level, and past knee injuries. Approximate ages can be assumed by the given ski racing level. I received 124 responses, and first grouped participants according to their racing competition level: U16, Pre Professional, and Professional. All U16s race in full-time academy programs in the United States Western, Rocky, or Eastern Divisions. All other athletes race under The Fédération Internationale de Ski (FIS).

### 2.2 Data analysis

I created a database for responses in Excel and used this platform to summarize results. Each data point reflects a single time an injury occurred, or that the participant has never injured a knee. I used variables including racing level, knee angle, as well as scores for tears of the ACL, PCL, MCL, LCL, patellar tendon, meniscus, and total tears. I have summarized this data and compared knee ligament injuries with Q-angles and ski racing ability. I used R-studio to run a one way analysis of variance (ANOVA) on the data [3]. This code also generated boxplots to visualize the data.

## 3. Results

Out of 124 participants, 57 participants tore their knee at least once in their career. Of the 124 surveys returned, there were 30 Professional, 62 Pre Professional, and 32 U16 participants. This means that of the survey participants, 45.16% of female U16 through Professional alpine ski racers have torn a knee at least once (Fig. 1). Of the Professional, Pre Professional, and U16 racers 83.33%, 27.42%, and 43.75% had torn a knee at least once, respectively (Fig. 1).

**Figure 1.**
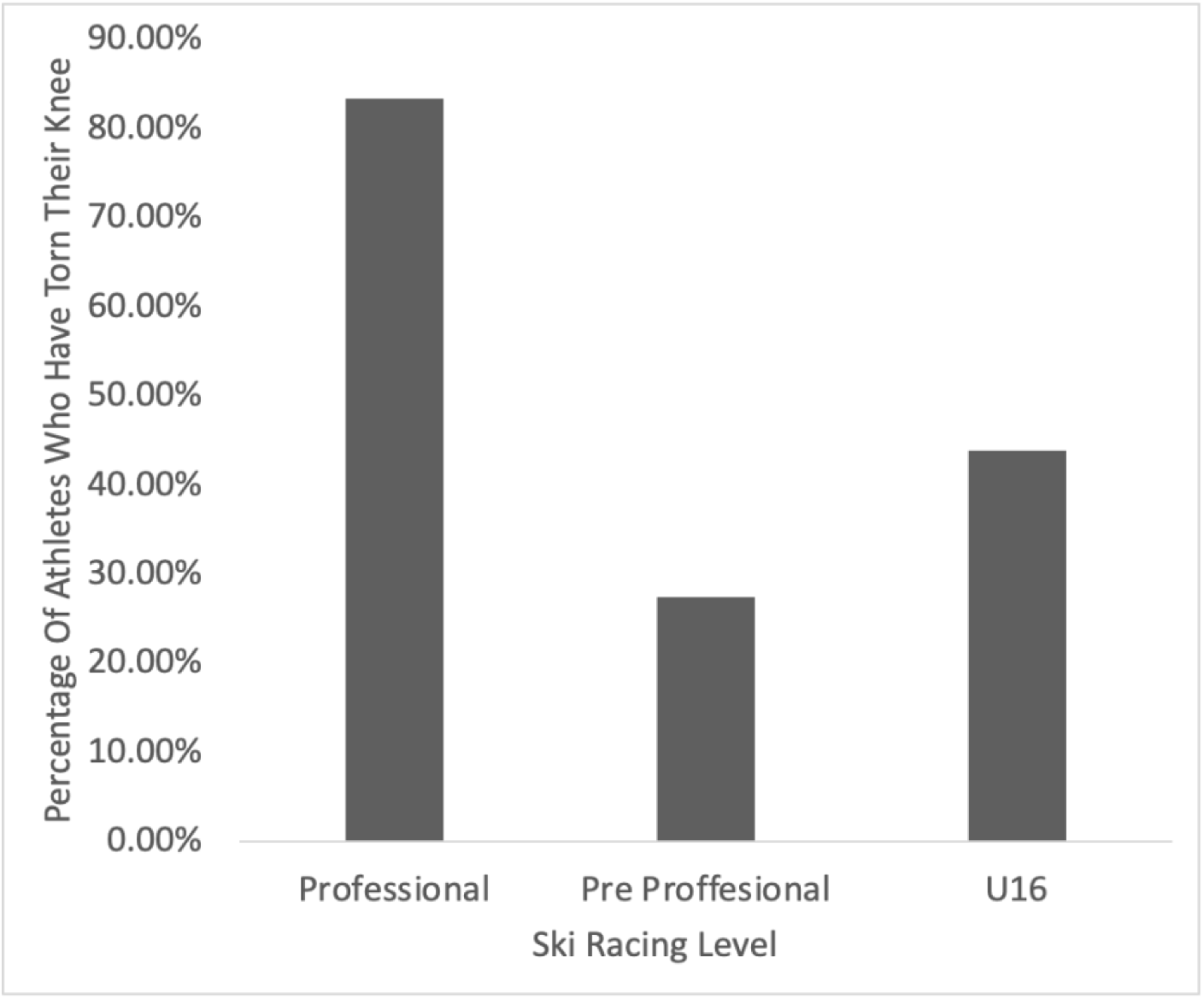
The percentage of athletes within each ski racing level who have torn their knee at least once (n = 124)

While 57 athletes reported at least one ligament tear, there was a total number of 190 tears reported. Athletes at the Professional level tore more than twice as many ligaments as the Pre Professional or U16 group, totaling 102 torn ligaments, making up 53.68% of the total ligaments torn (Fig. 2). The average number of tears among skiers at the Professional level was 3.34 (Fig. 3). Skiers of the Pre Professional group tore a total of 48 ligaments averaging 0.81 ligaments, and the U16 group tore a total of 40 ligaments averaging 1.25 ligaments (Fig. 2). The number of torn ligaments within the Professional group compared to the Pre Professional and U16 groupings were both statistically significant, *P<* 0.0001 and *P*= 0.002, respectively (Fig. 3). The difference in the number of torn ligaments among skiers at the Pre Professional and U16 groupings was not statistically significant, *P* = 0.4371 (Fig. 3).

**Figure 2.**
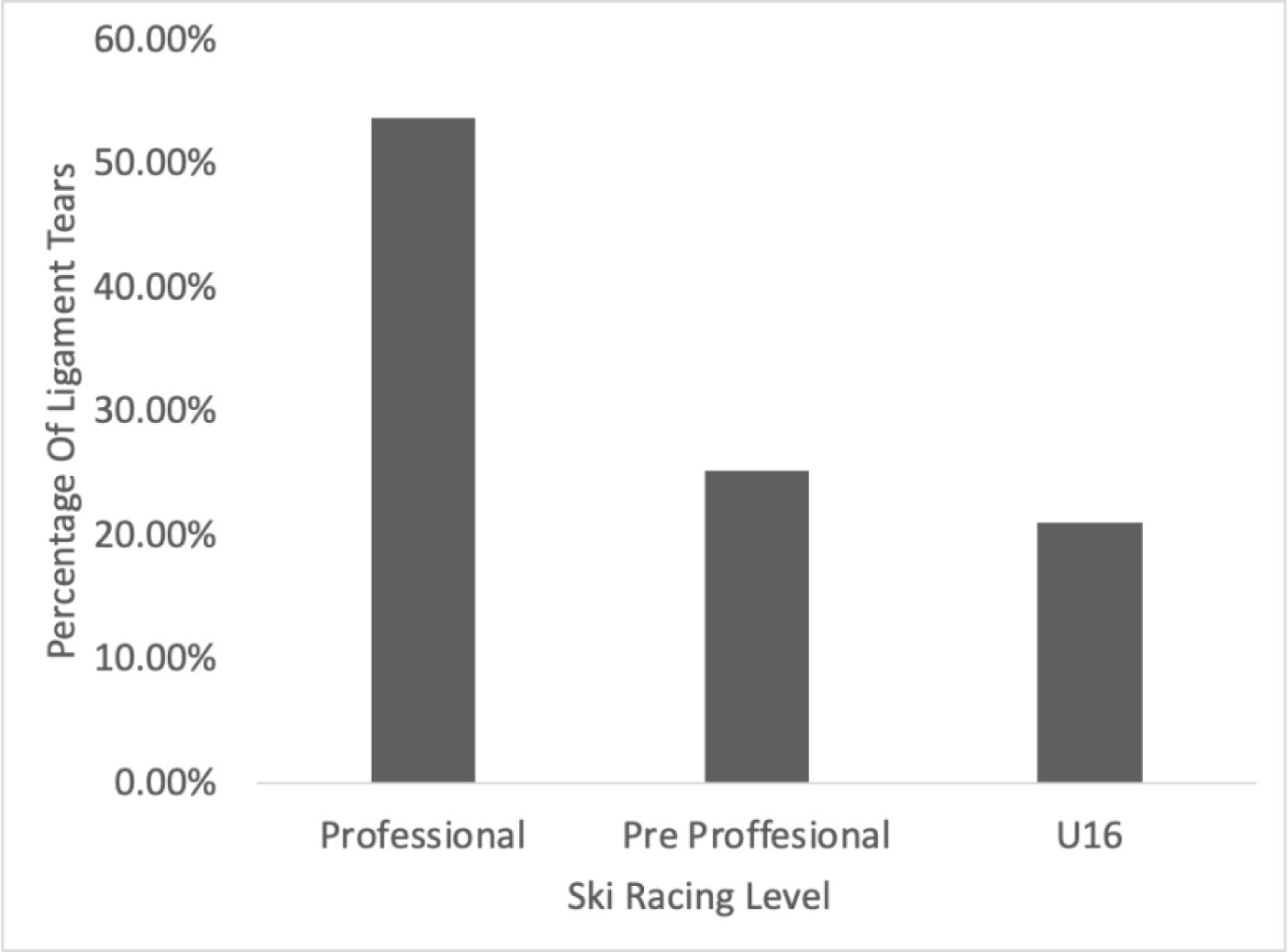
The percentage of ligament tears within each ski racing level (n=190)

**Figure 3.**
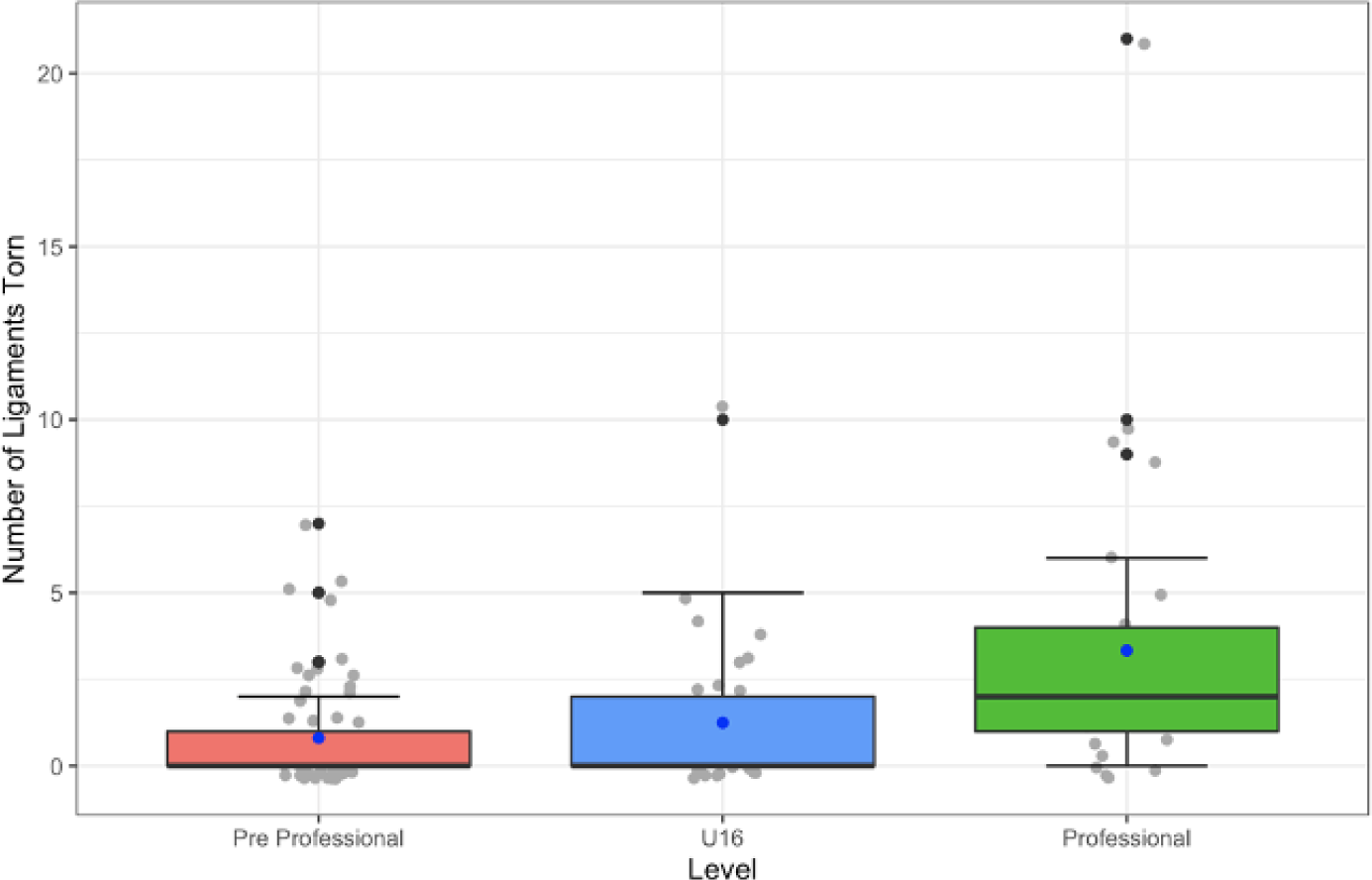
The number of ligament tears within each level of ski racing (n=124), represented by boxplots, dotplots, and standard error of the mean (SEM) plot. Gray dots=sample data points, black dots=outliers, and blue dots=means

The greatest number of knee injury incidents occurred among skiers at the Professional level, totaling 46.32% of the total injury incidents and averaged 1.47 incidents (Figure 4). An incident counts as a singular injury occurrence, where at least one ligament was torn. There were about twice as many injury incidents among skiers at the Professional level as Pre Professional or U16 level. Skiers at the Pre Professional and U16 level had 29.32% and 24.21% of injury incidents, averaging 0.45 and 0.72 incidents, respectively (Figure 4). There was no significant difference in the number of injury incidents among skiers at the Pre Professional level and U16 level (*P* = 0.223, Figure 5). The difference in the number of injury incidents between skiers at the Professional level compared to those at the Pre Professional and U16 levels were both statistically significant (*P* < 0.0001 and *P* = 0.0039, respectively, Figure 5). Overall, the number of injury incidents and the number of ligaments torn increased as the ski racing level increased.

**Figure 4.**
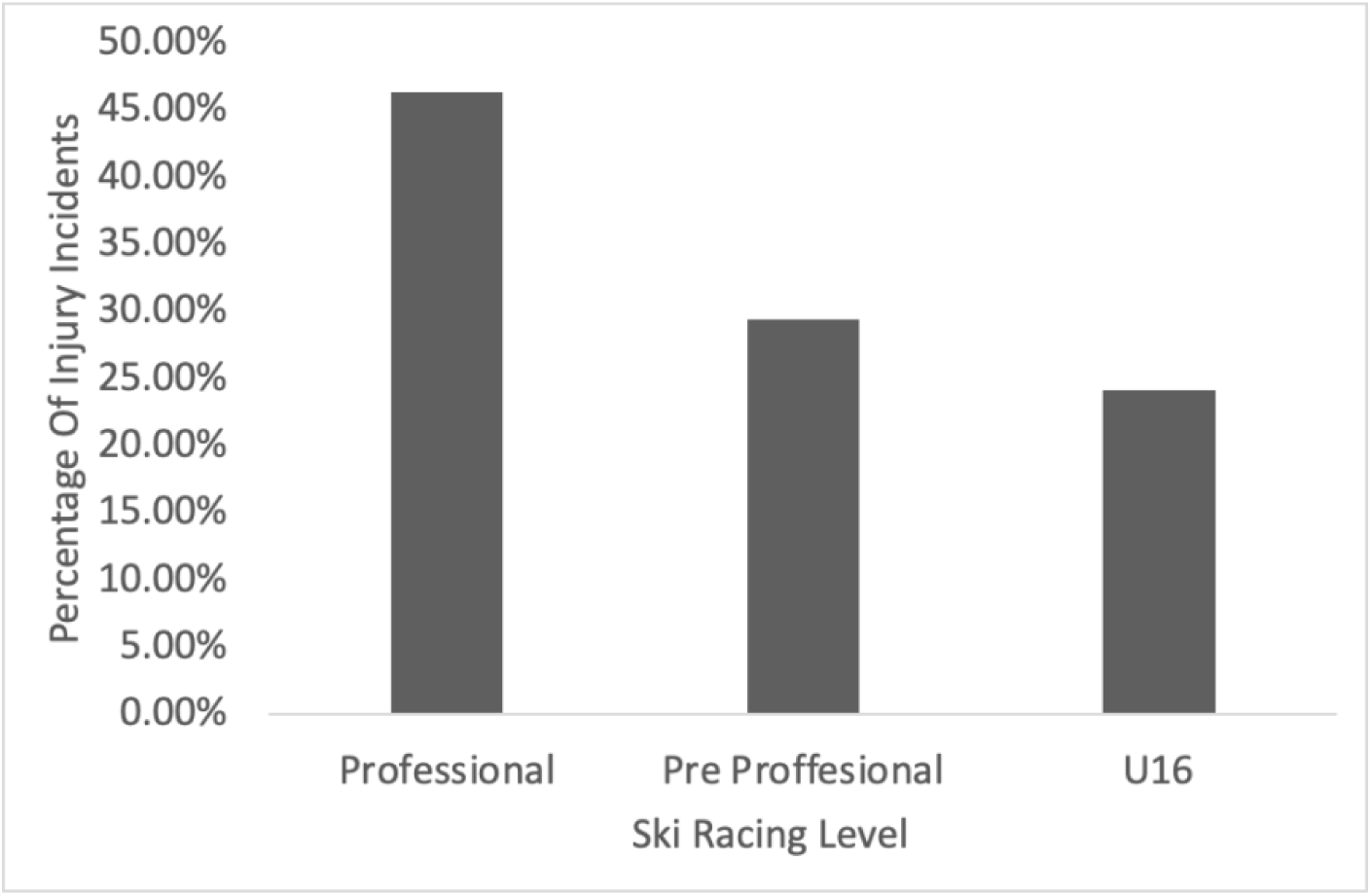
The frequency of knee injuries incidents within each level of ski racing (n=95)

**Figure 5.**
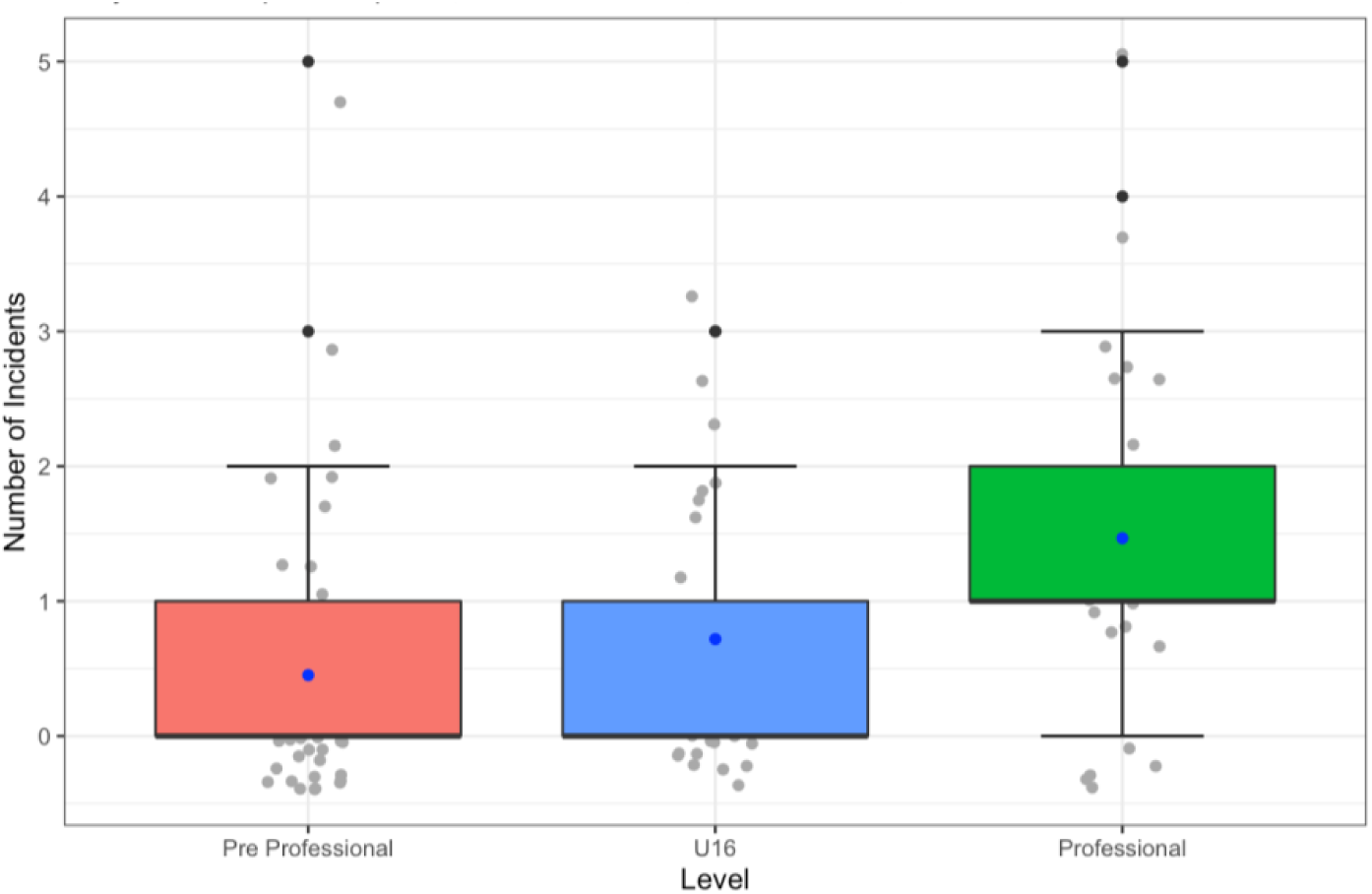
The number of injury incidents at each ski racing level (n=124) represented as boxplot, dotplot, and standard error of the mean (SEM) plots. Gray dots=sample data points, black dots=outliers, and blue dots=means

The majority (51.61%) of participants presented with a Q-angle of about 15°, 19.35% presented with a Q-angle of about 20°, and 7.26% presented with a Q-angle of about 10° (Figure 6). There was a grouping of 21.77% that were unsure of their relative Q-angle which comprised the N/A group (Figure 6). Over half of the athletes with about a 15° and about a 20° Q-angle 54.69% and 54.17%, respectively, have torn a knee at least once (Figure 7). Athletes with about a 10° Q-angle also had a high percentage of torn knees, with 44.44% of athletes tearing a knee at least once. However, only 22.22% of the N/A group has torn a knee at least once, which is much less than any of the other groupings (Figure 7).

**Figure 6.**
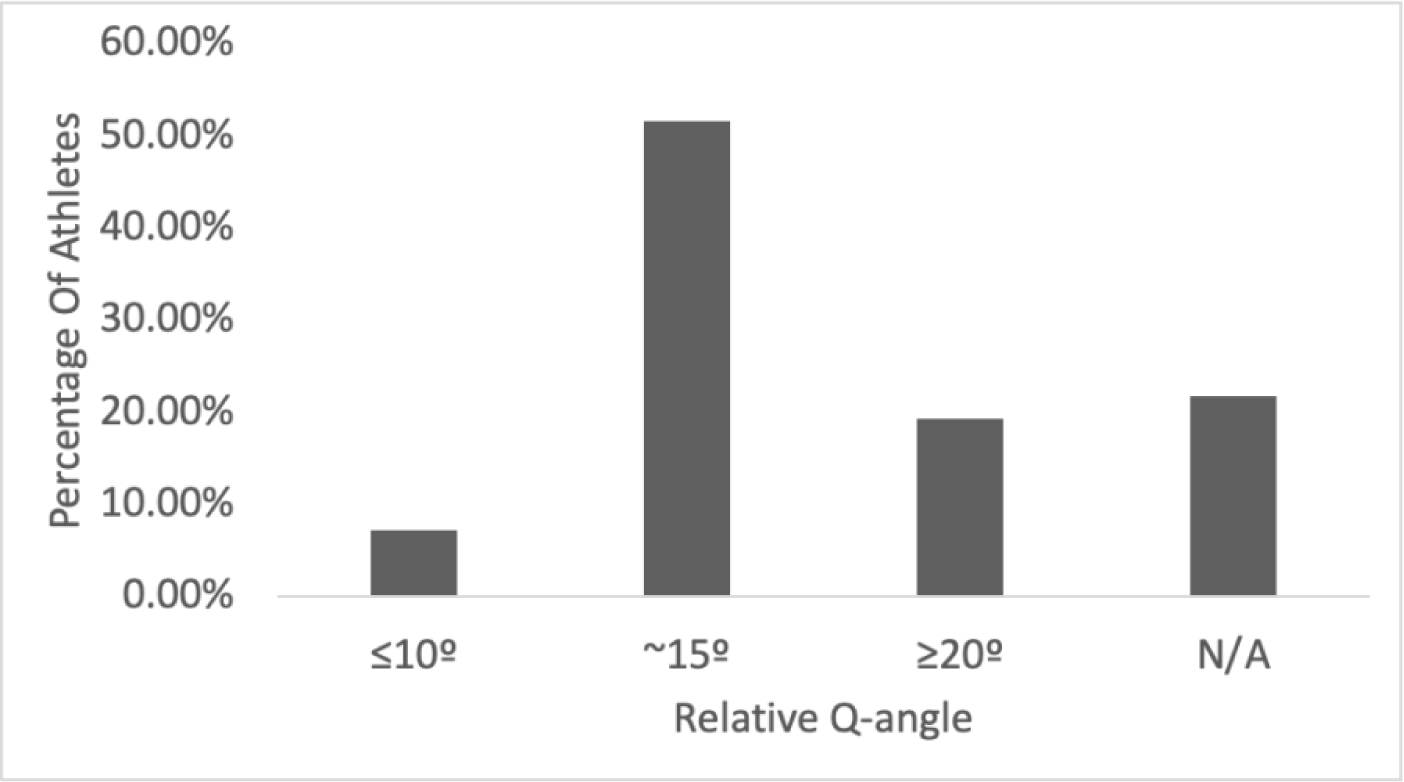
Percentage of athletes presenting with each relative Q-angle (n=124)

**Figure 7.**
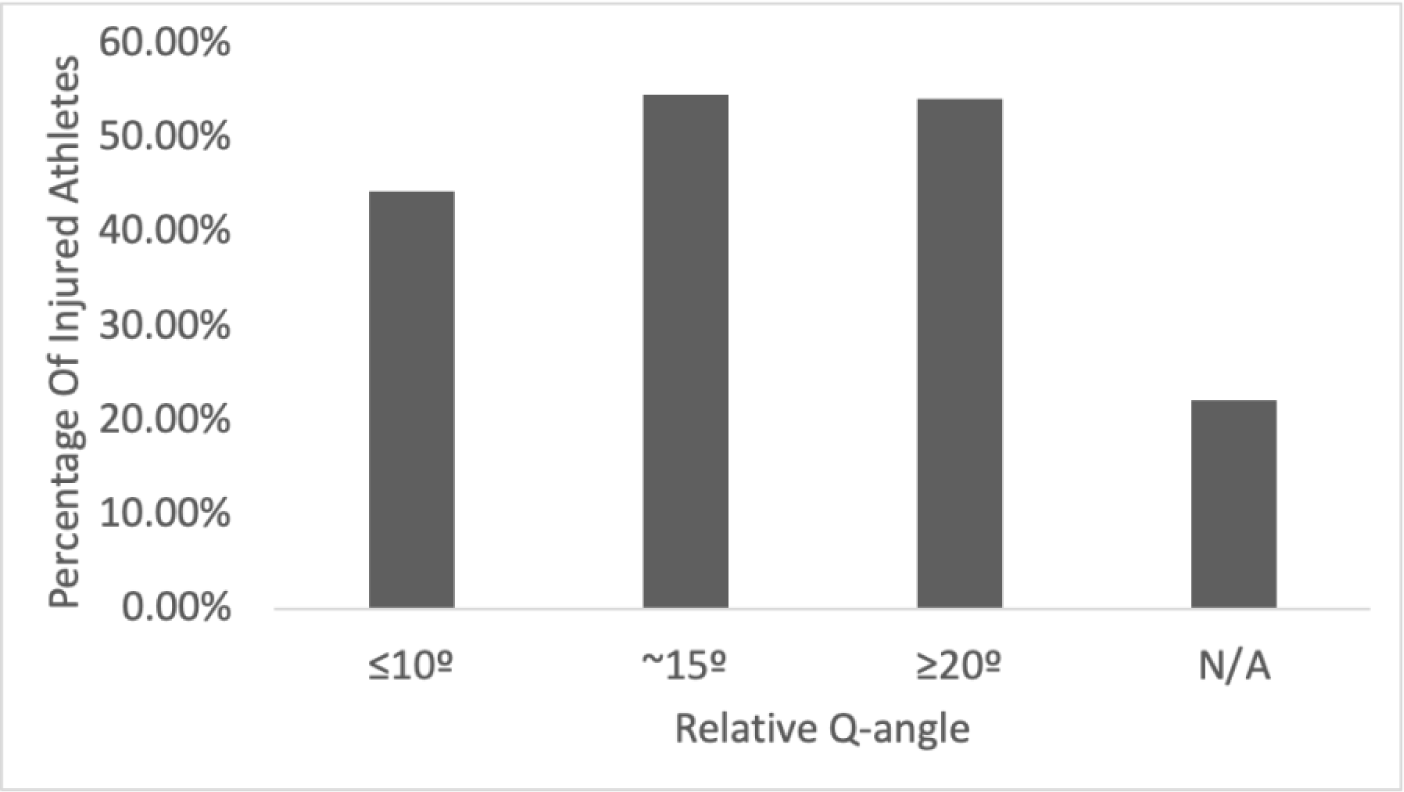
Percentage of athletes with each relative Q-angle who have experienced a knee ligament tear at least once (n=124).

The greatest number of injury incidents occurred within athletes with about a 15° Q-angle (57.89%), which is about twice as many injury incidents as athletes with about a 20° Q-angle experienced, averaging 0.89 incidents with a P-value of 0.4363≥0.05 (Figure 8). Athletes with about a 20° Q-angle experience 30.53% of the injury incidents. Injury incidents occurring within the N/A and 10° Q-angle groupings were very small percentages of the total injury incidents that occurred, 3.16% averaging 0.125 incidents and 8.42% averaging 0.89 incidents (Figure 8). There were only two significant differences in the number of injury incidents when Q-angle categories were compared: skiers with Q-angles of 20° with the N/A grouping and of 15° with the N/A grouping (*P* = 0.0019 and *P* = 0.003, respectively, Figure 8).

**Figure 8.**
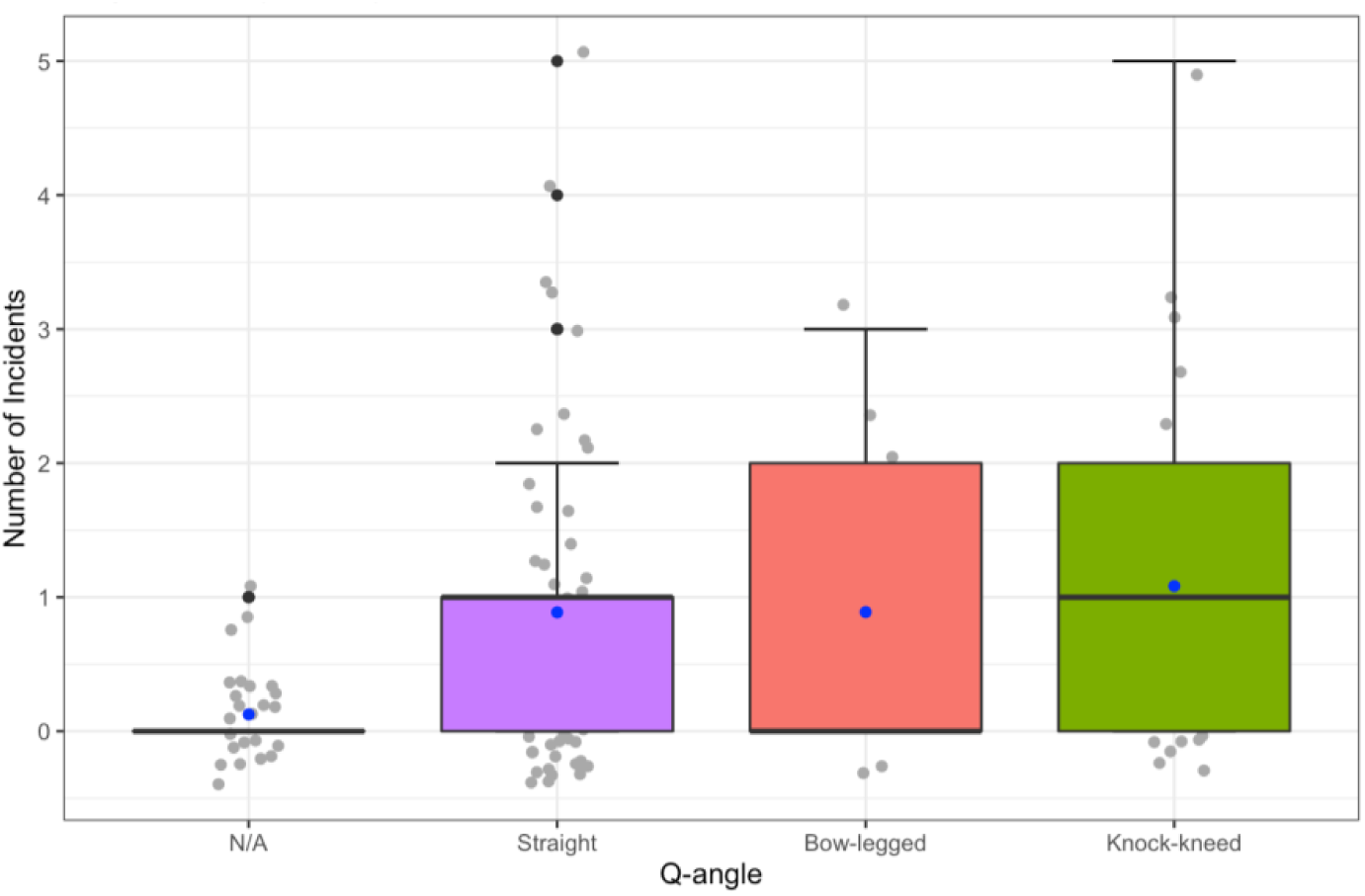
The number of injury incidents compared to the distribution of general Q-angles (n=95) represented as boxplot, dotplot, and standard error of the mean (SEM) plot. Gray dots=sample data points, black dots=outliers, and blue dots=means.

Most ligament tears occurred in athletes with about a 15° or 20° Q-angle, accounting for 84.21% of the total ligaments torn (Figure 7). Athletes with a 15° Q-angle tore the most ligaments (57.89%), averaging 1.77 tears (Figure 9). Athletes with a 20° Q-angle tore about half as many (26.32%) total ligaments than those with a 15° Q-angle, though this difference was not statistically significant (*P* = 0.644). However, athletes with a 20° and 10° Q-angle averaged the highest number of ligament tears per incident, 2.08 and 2.22, respectively, and this difference was also not statistically significant (*P*= 0.898, Figure 9). The difference in the number of injury incidents for skiers with Q-angles of about 10° and Q-angles of about 15° was also not statistically significant (*P* = 0.651, Figure 9).

**Figure 9.**
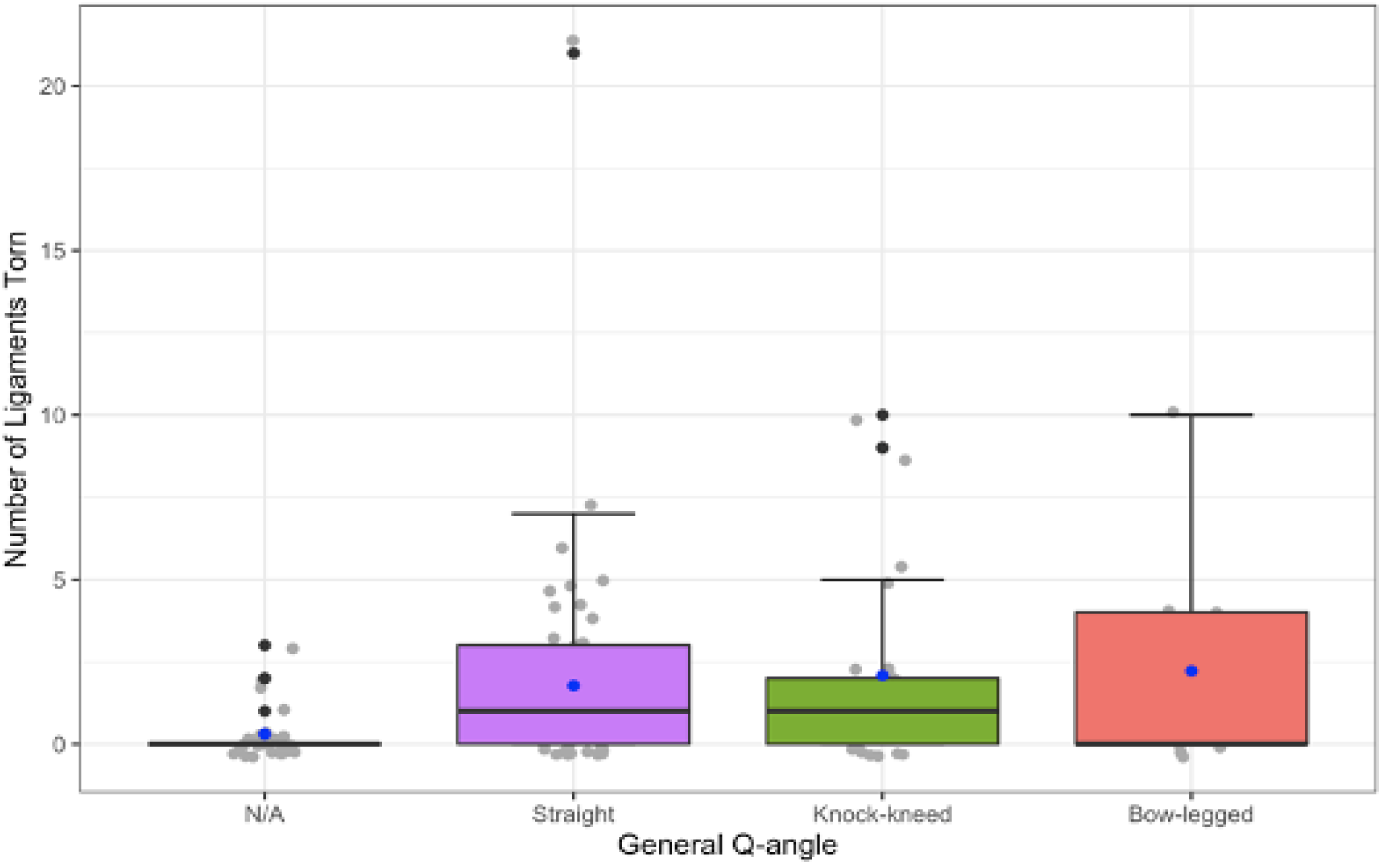
The number of ligament tears within each general Q-angle grouping (n=190) represented by boxplot, dotplot, standard error of the mean (SEM) plots. Gray dots=sample data points, black dots=outliers, and blue dots=means.

The ACL, MCL, and meniscus were the most frequently torn and represented 89.56% of the total ligament tears (Figure 10). The ACL made up 35.16% of the total tears, 21.43% were the MCL, and 32.97% were the meniscus. The remaining tears were split between the PCL, LCL, and patellar tendon totaling 2.11%, 4.74%, and 3.68%, respectively (Figure 10). The ACL, MCL, and Meniscus were the most commonly torn as well as the most frequently torn in athletes presenting with about a 15° or 20° Q-angle (Figure 11).

**Figure 10.**
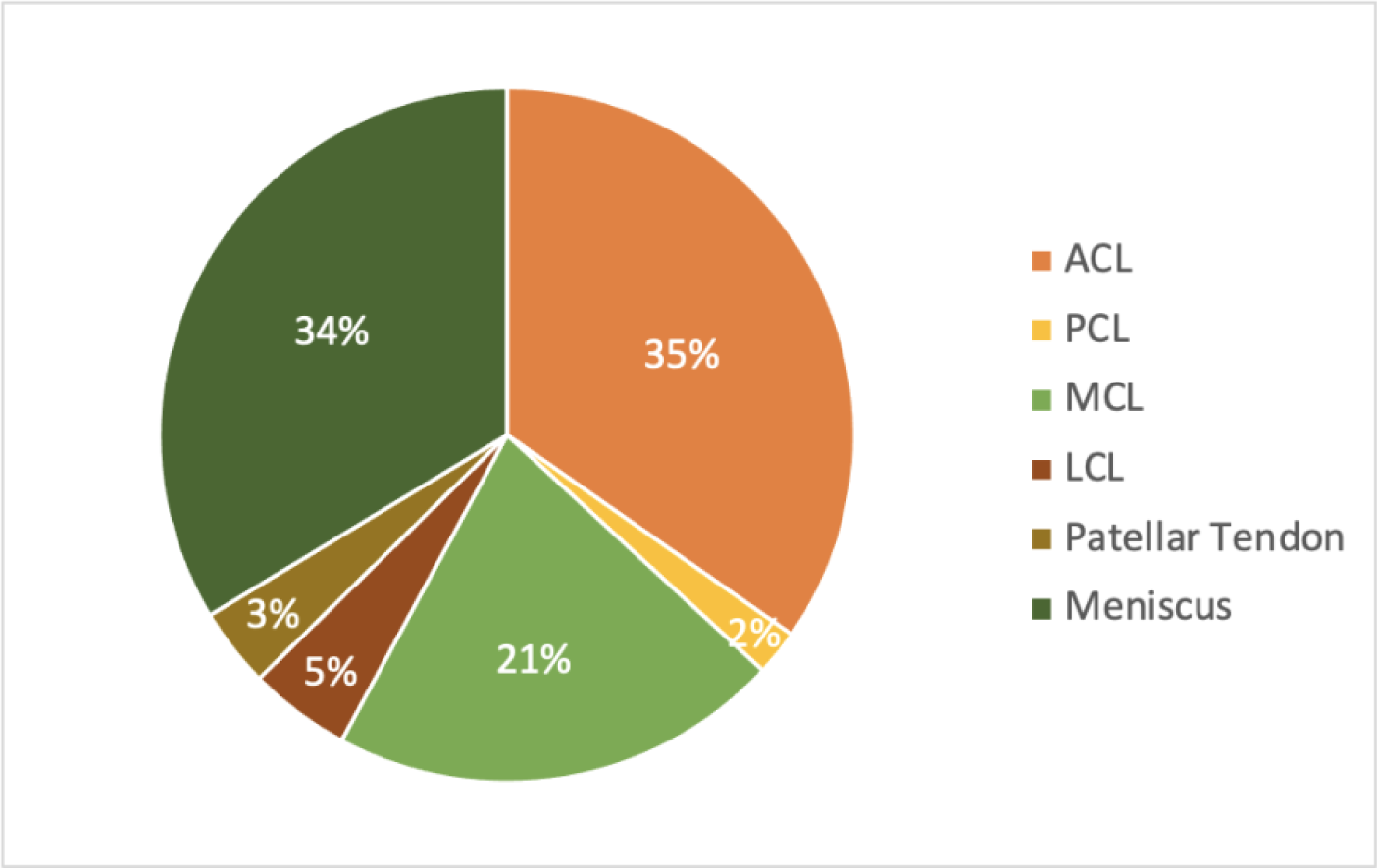
The percentages of all ligaments of the knee that had been reported torn by alpine ski racers (n= 190)

**Figure 11.**
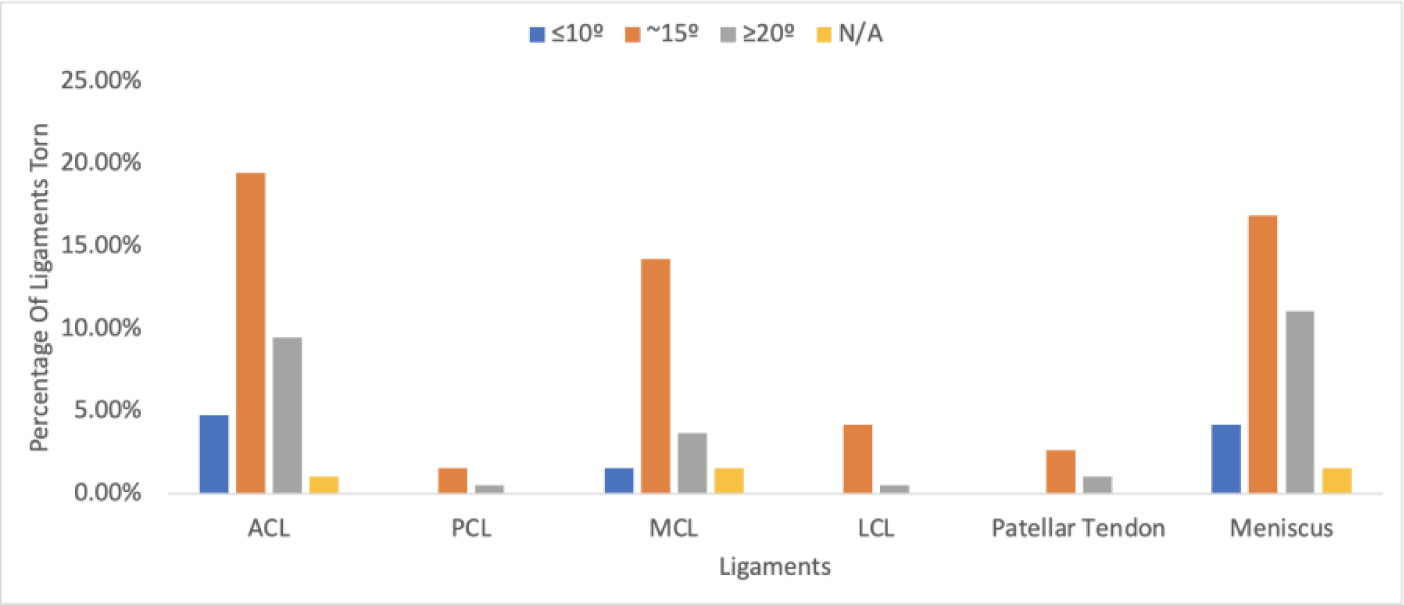
Percentage of each ligament torn broken down by relative Q-angles (n=190)

Athletes presenting with about a 15° Q-angle were most susceptible to ACL and MCL tears, totaling 13.84% and 11.11%, respectively. Athletes presenting with about a 20° Q-angle were most susceptible to meniscus tears totaling 11.11% (Figure 12). Athletes in the N/A grouping and athletes presenting with about a 10° Q-angle were not particularly susceptible to any specific ligament injury. Not only were the ACL, MCL, and meniscus the most frequently torn, but they were often torn concurrently. Combination tears of the ACL and meniscus are 18.51% of the total 108 tears involving the ACL, MCL, and/or meniscus (Figure 12). Combination tears of the ACL, MCL, and meniscus comprise a total of 14.81% while combination tears of the MCL and Meniscus are only 6.47% (Figure 12). The least frequented combination tear was of the ACL and MCL which only occurred in athletes with a Q-angle of about 15° or 20° totaling 1.84% (Figure 12).

**Figure 12.**
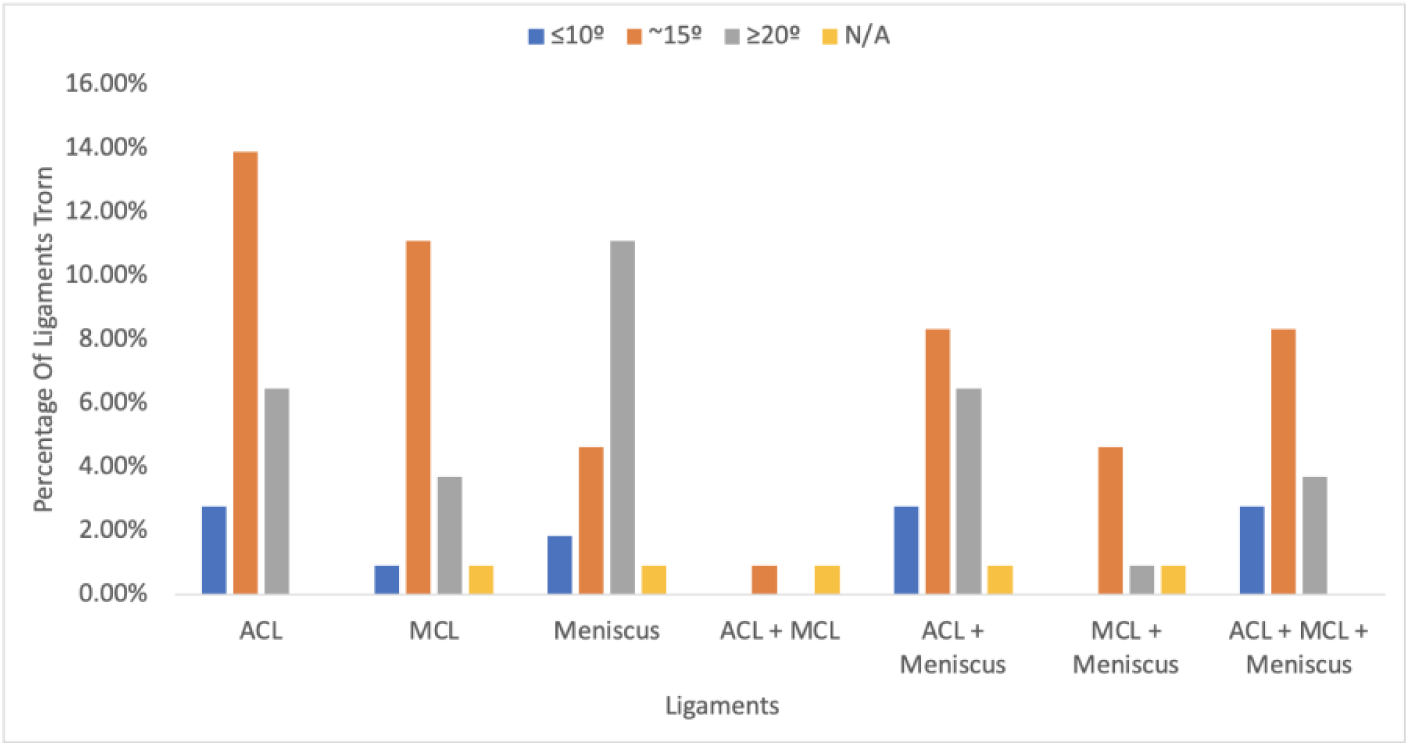
The percentage of tears including the ACL, MCL, and/or meniscus broken down by ligaments and combination groupings differentiated by the relative Q-angles (n=105)

Many athletes (18.55%) tore their knee more than once thus far in their career (Figure 13). The maximum number of injury incidents sustained by a singular athlete was five, but this occurred only within the Professional and Pre Professional level. Athletes within the U16 level sustained no more than three injury incidents (Figure 13). Many of these injury incidents were re-tears of a ligament. Re-tears occurred in the ACL, MCL, LCL, and meniscus totaling 28.42% of the total ligaments torn (Figure 14). All the LCLs torn were re-torn, while about a third of the ACLs, MCLs, and meniscus were re-torn (Figure 14). No PCLs or patellar tendons were re-torn. Most of the re-tears, 61.11%, occurred at the Professional level with still a large percentage, 20.37%, of re-tears occurring at the Pre Professional level (Figure 15). Most re-tears occurred in athletes with larger Q-angles of 15° or 20° totaling 87.04% of the ligaments torn a second time (Figure 16). All the LCL tears, and most of the MCL tears (91.67%) occurred in athletes with about a 15° Q-angle (Figure 16)

**Figure 13.**
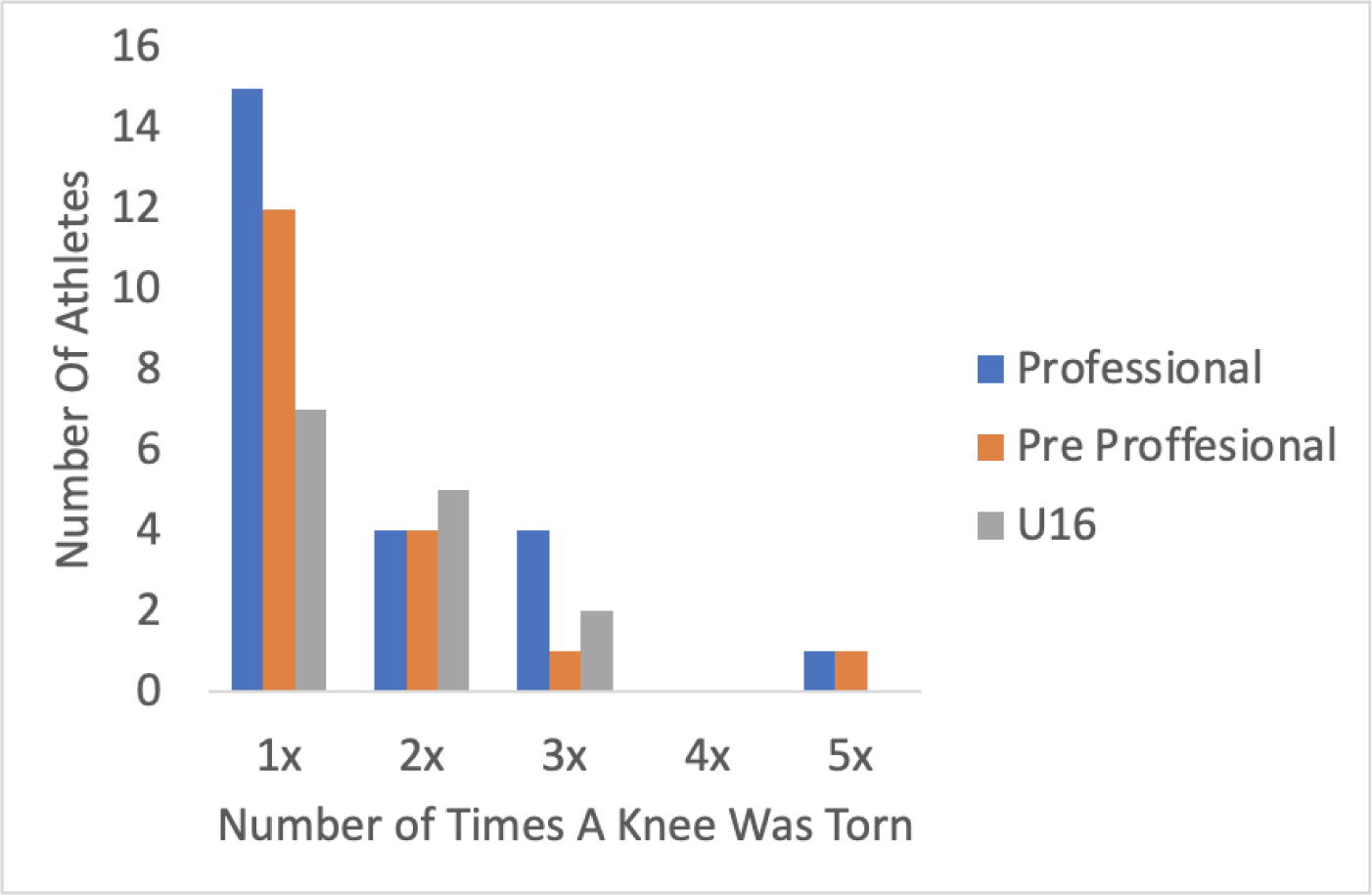
The number of times participants have torn their knee, categorized by their ski racing level (n = 95)

**Figure 14.**
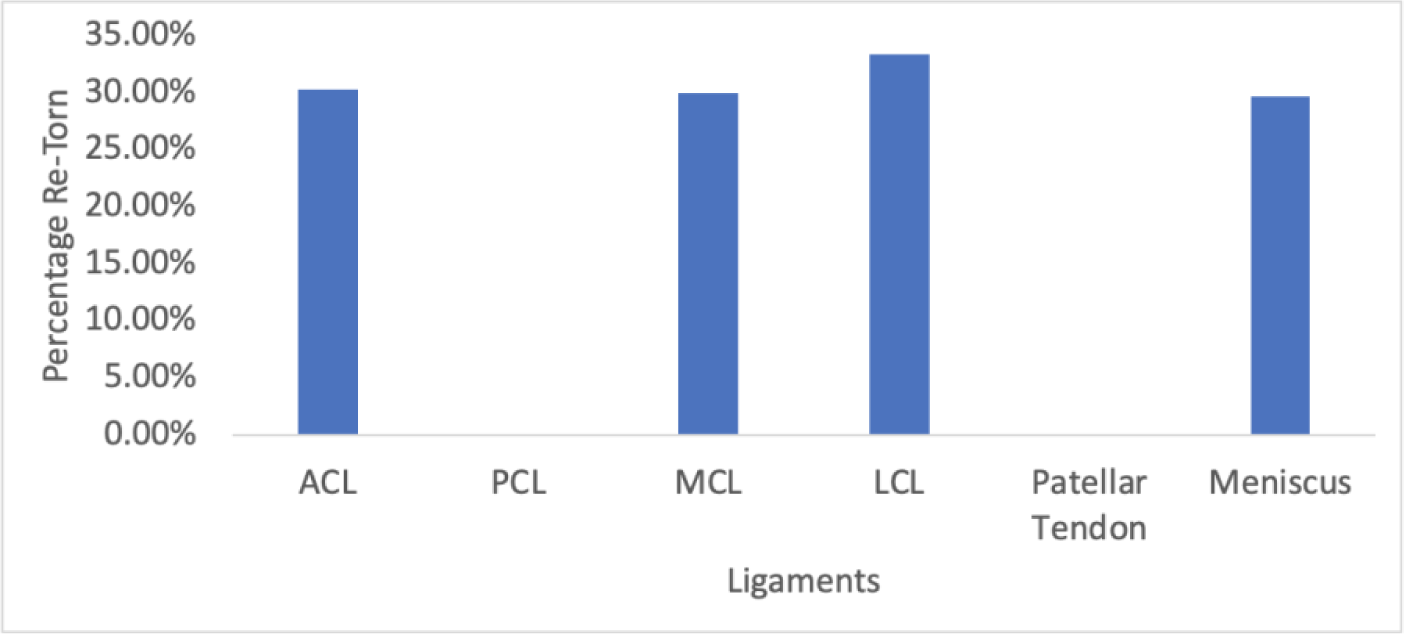
Percentage of each ligament re-torn (n=190)

**Figure 15.**
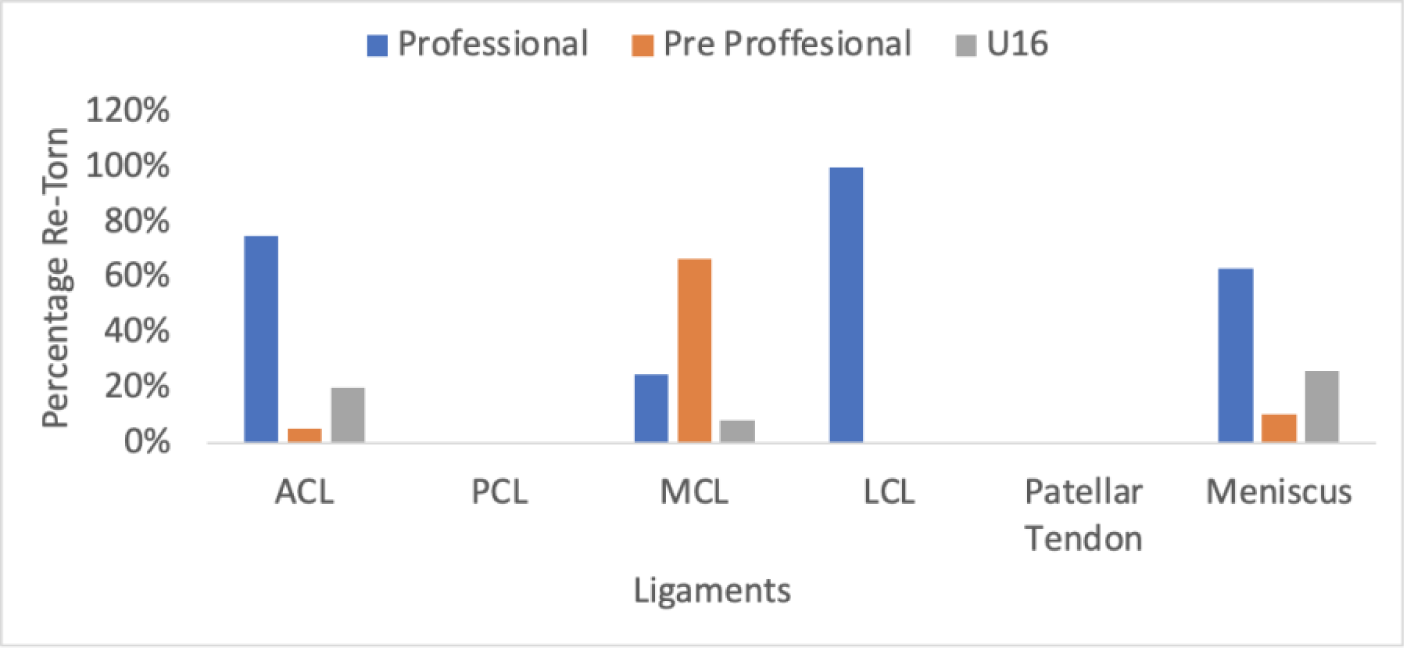
Percentage of each ligament re-torn differentiated by ski racing level (n=54)

**Figure 16.**
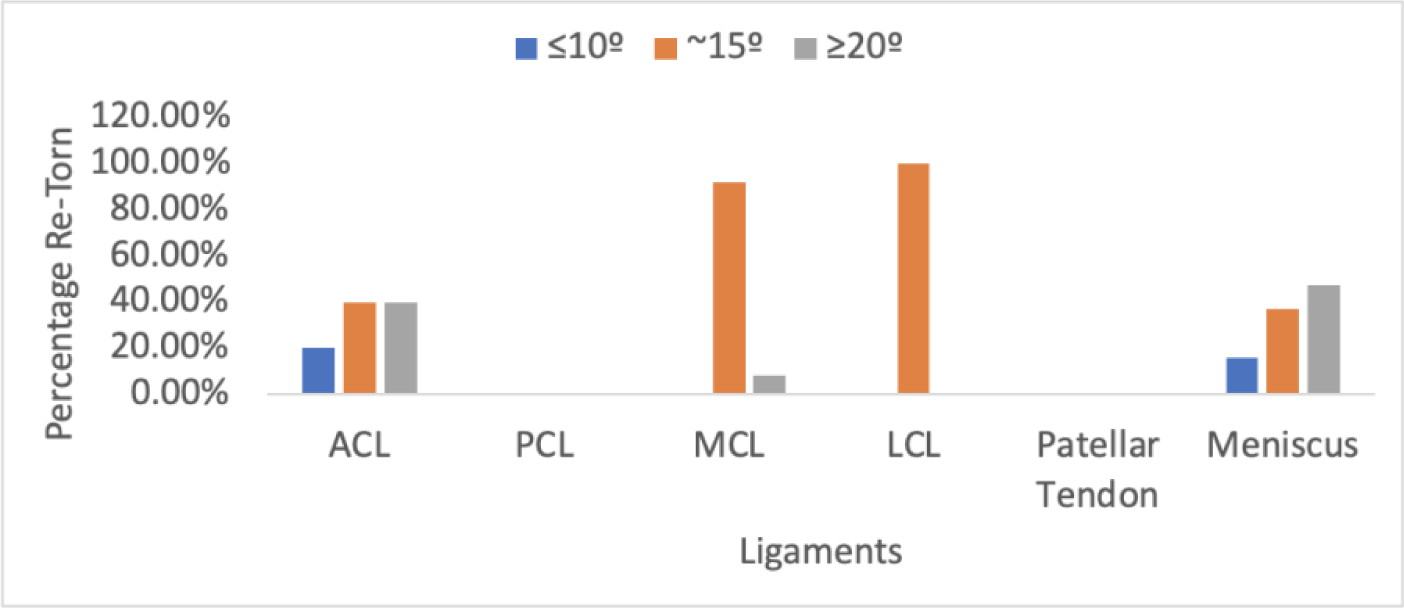
Percentage of each ligament re-torn differentiated by relative Q-angles (n=54)

## 4. Discussion

Results support the hypothesis that higher levels of ski racing led to a high risk of knee injuries in participating athletes. Almost half (45.16%) of the participants had torn a knee ligament at least once thus far in their career. A total of 83.33% of the Professional athletes had torn a knee ligament at least once which is 3.04 times more than athletes of the Pre Professional group and 1.90 times more than the U16 athletes (Figure 1). The risk of knee injury increases as ski racing competition level increases, with the most injuries occurring at the Professional level. Conditions are relatively similar for the U16 and Pre Professional levels, the main difference being that most U16 athletes are just beginning adolescent development, so their Q-angles are typically relatively smaller, potentially resulting in less injuries. The Professional athletes typically can produce more power when skiing therefore combating greater forces, so it makes sense that their injury rate would be so much higher than those of the Pre Professional and U16 athletes.

Athletes at the Professional level tore 102 total ligaments with an average of 3.33 ligaments per injury incident. This is twice the number of ligaments torn per injury incident than at the Pre Professional or U16 levels (Figure 2). The increase in tears from the Pre Professional and U16 groups to the Professional group is significant and shows that the risk of injury is much higher at the Professional level. This would indicate that the forces that occur during crashes at the Professional level are much greater than that at the Pre Professional or U16 level. Bode Miller, a Professional racer, has been clocked at 12 gravitational forces (Gs) within the course; 12Gs is the same level of force that fighter-jet pilots experience [1]. Crashing with this amount of force would explain why the injury rate at the Professional level is so high.

Q-angles also play a role in the risk of knee injuries for female alpine ski racers. My hypothesis about how the Q-angle will influence knee injuries was partially supported. Injuries occurring in athletes with a 15° or 20° Q-angle made up 85.26% of the total ligaments torn showing that larger Q-angles were much more susceptible to knee injuries, but these differences were not statistically significant (Figure 8). A larger data set would be useful for differentiating significant differences in Q-angles.

I predicted that a Q-angle of 20° renders an athlete to be more prone to injury of the MCL, ACL, and patellar tendon and results partially support this. There were very few patellar tendon tears, but all the tears occurred in athletes with a 15° or 20° Q-angle (Figure 12). Individually, the ACL and MCL are most susceptible to injury in athletes with a Q-angle of 15°, but not 20°. The average female, athlete or not, has a Q-angle of 15°, meaning that in general female ski racers are mainly at risk for ACL and MCL tears. I also predicted that an athlete with a Q-angle of 15° would be more prone to injury of the ACL and meniscus, however, the meniscus was most susceptible to injury in athletes with a Q-angle of 20°. Combination tears of the ACL, MCL, and meniscus were also very frequent in athletes with a Q-angle of 15° and 20° (Figure 12). A larger data set would be beneficial for providing further results on combination tears.

Finally, I predicted that an athlete with a Q-angle of 10° would be more prone to injury of the LCL, which was not supported by my data. All the LCL tears occurred in athletes exhibiting a Q-angle of 15° or 20° (Figure 12). The most frequently torn ligament for athletes with a Q-angle of 10° is the ACL, but these tears account for only 2.78% of the total ACL tears that occurred. There were fewer athletes who reported having a Q-angle of 10° than 15° or 20°, so more data are needed to make conclusions about injuries in female athletes with Q-angles of about 10°. However, this discrepancy is consistent with the fact that females generally have much larger Q-angles than men.

There were only four PCL tears with each occurring in an athlete exhibiting a Q-angle of about 15° or 20°, and one athlete having two of the PCL tears. This data set is too small to make an accurate assumption about which Q-angle influences the likelihood of PCL tears. However, half of the PCL tears occurred at the Professional level and the other half occurred at the Pre Professional level, and none of the U16 athletes reported this injury. This is consistent with my hypothesis that the higher the level of ski racing one is participating in, the more likely it is for them to tear their knee. PCL tears most commonly occur during car crashes, so the forces needed to tear are extremely large and are probably much greater than what most U16 athletes would experience [2].

The likelihood of athletes having a knee injury more than one time was also greatest at the Professional level, with 61.11% of the re-tears occurring at this level (Figure 15). This is three times more than at the Pre Professional level and 3.3 times more than at the U16 level. It is consistent that re-tears are also more prevalent in the high levels of racing, however there were about the same number of re-tears at the Pre Professional and U16 levels.

Overall females with larger Q-angles were more likely to tear their knee in some capacity than females with smaller Q-angles. Having a larger Q-angle puts more continual stress on the ligaments of the knee which is why athletes exhibiting larger Q-angles are more likely to tear their knee than those with smaller Q-angles, but more research is needed to figure out how to prevent female ski racers from enduring so many knee injuries. Injuries and the number of ligaments torn during each injury incident are more likely to occur at higher levels of ski racing. Combined, Professional ski racers with larger Q-angles are the most at risk for injury.

Jeannie Thoren, a member of the U.S. Ski and Snowboard Hall of Fame, has been researching equipment adjustments that can be made to female alpine ski racing equipment to help improve ability and decrease injuries. Alpine racing gear is not gender specific, so men and women use the same equipment except for the length of skis allowed by FIS. It is much more difficult for a woman to “get forward” on her skis, a problem that worsens on steeper slopes because of greater Q-angles. Thoren has been experimenting with adding heel lifts to ski boots to lift the pelvis forward, moving the center of gravity forward, and creating a more natural forward stance [6]. However, this cannot be implemented at the Pre Professional or Professional level because of FIS regulations surrounding Boot Height, which is defined as the height from the sole to the top of the footbed. FIS requires this height to be 43mm or less, which does not allow for room to experiment with lifting the heel of the footbed. Experimentation with female oriented equipment changes could allow for a much lower injury rate as well as potentially higher performance from female athletes at all competition levels.

Female ski racers have a high risk of knee injury, and this risk increases with Q-angle size as well as ski racing level. This problem needs to be addressed by potential equipment modifications, such as those suggested by Jeanie Thoren, and by more research. Further research should include exact measurements of the Q-angle as well as a larger data set. Equipment changes may need to be different for U16 athletes compared to Pre Professional and Professional athletes because of the different stages of anatomical development they are undergoing. This study shows that ski racing level and Q-angle metrics play a significant role in the likelihood of knee injuries.

## Data Availability

All data produce in the present study are available upon reasonable request to the authors

## Notes

### Competing Interest Statement

The authors have declared no competing interest.

### Funding Statement

This study did not receive any funding

### Author Declarations

IRB of Colby College gave ethical approval for this work

